# Natural language inference for clinical registry curation

**DOI:** 10.1101/2021.06.14.21258493

**Authors:** Bethany Percha, Kereeti Pisapati, Cynthia Gao, Hank Schmidt

## Abstract

Clinical registries - structured databases of demographic, diagnosis, and treatment information for patients with specific diseases or phenotypes - play vital roles in high-quality retrospective studies, operational planning, and assessment of patient eligibility for research, including clinical trials. However, registries are extremely time and resource intensive to curate. Natural language processing (NLP) can help, but standard NLP methods require specially annotated training sets or the construction of separate models for each of dozens or hundreds of different registry fields, rendering them insufficient for registry curation at scale. Natural language inference (NLI), a specific branch of NLP focused on logical relationships between statements, presents a possible solution, but NLI methods are largely unexplored in the clinical domain outside the realm of conference shared tasks and computer science benchmarks. Here we convert registry curation into an NLI problem, applying five state-of-the-art, pretrained, deep learning based NLI models to clinical, laboratory, and pathology notes to infer information about 43 different breast oncology registry fields. We evaluate the models’ inferences against a manually curated, 7439 patient breast oncology research database. The NLI models show considerable variation in performance, both within and across registry fields. One model, ALBERT, outperforms the others (BART, RoBERTa, XLNet, and ELECTRA) on 22 out of 43 fields. A detailed error analysis reveals that incorrect inferences primarily arise through models’ misinterpretations of temporality--they interpret historical findings as current and vice versa--as well as confusion based on subtle terminology and abbreviation variants common in clinical notes. However, modern NLI methods show promise for increasing the efficiency of registry curation, even when used “out of the box” with no additional training.

## Introduction

Academic medical centers, pharmaceutical companies, and other healthcare organizations routinely develop clinical registries: structured databases of demographic, treatment, and disease-related information for patients with specific diseases or phenotypes. The creation and maintenance of these registries has traditionally relied upon human curators, who sift through thousands of clinical documents, manually abstracting relevant information into database software **[1]**. The structured data they create form the foundation of high-quality retrospective analyses and ongoing assessment of patient eligibility for research studies, including clinical trials **[2]**. Registries also play an important role in generating “real world evidence” for the pharmaceutical regulatory approval process **[3]**. Manual registry curation, however, is expensive, time-consuming, and prone to human error. As registries grow, the time and effort required to enter new patient information compete with a constant need to update the records of existing patients, causing curators to become bottlenecks and delaying registries relative to the true state of patient care. These delays, in turn, create larger delays in upstream registries that draw data from multiple institutions **[4, 5]**.

Given these challenges, the development of natural language processing (NLP) methods that automatically extract and structure registry information from text would have profound implications, both for clinical research and patient care **[6]**. However, the task is extremely challenging: information pertaining to dozens of different registry fields must be extracted simultaneously, and patients’ conditions and treatment status both change over time. The most obvious NLP approach--building a separate text classification model for each registry field **[20]**--would necessitate the development of dozens or even hundreds of different models, each requiring separate annotated data for training. Indeed, any approach requiring annotated training data is likely to fail for one simple, practical reason: instead of hiring annotators, an institution could just as easily hire curators to build the registry itself.

The past few years have seen a reinvention of NLP methods around a class of large language models based on a novel neural network architecture called the transformer **[7]**. These models typically work in two stages. They are first “pretrained” using simple tasks like masked-word prediction, learning to model the regularities of language by repeating this task millions of times on massive, unlabeled corpora. The pretrained models are then “fine-tuned” to perform a variety of different NLP tasks. Most recently, these models have been applied to the famously difficult task of natural language inference (NLI), the task of deciding whether a given statement (the “hypothesis”) is implied to be true or false by another (the “premise”) **(Figure 1) [8]**. Also called “entailment recognition”, NLI closely resembles the task human curators perform in registry curation: reading the thousands of sentences that comprise a patient’s medical record, then deciding whether a series of other statements--corresponding to the structured data fields of the registry--are true. The most important feature of these models is their breadth: the same model can be applied to a potentially infinite number of different statement pairs. One well-performing NLI model could, therefore, be used to curate information for dozens or even hundreds of different registry fields.

**Figure 1:**
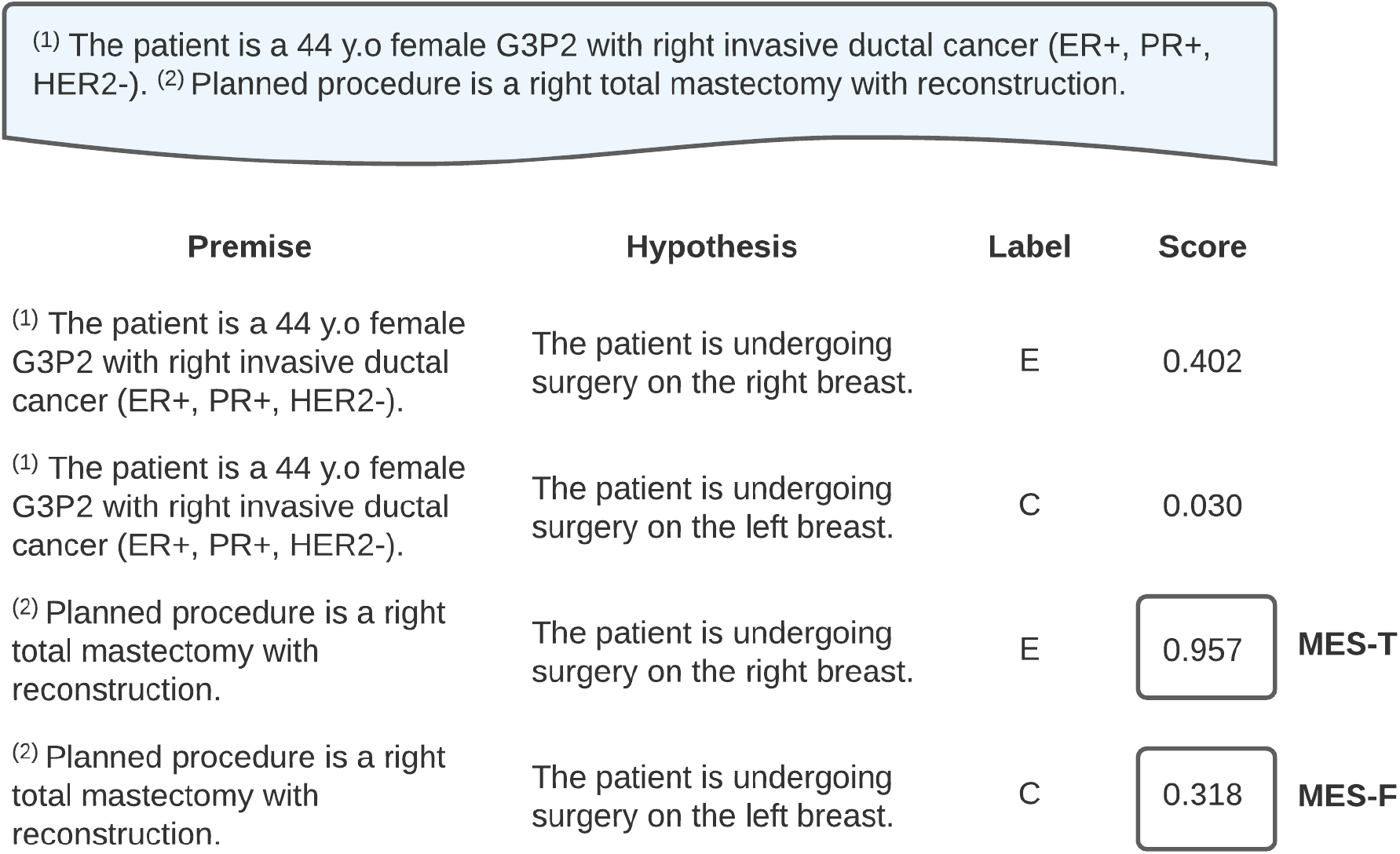
Natural language inference (NLI) model evaluation. An NLI model considers each premise sentence from a patient’s record (shaded blue) and compares it to a series of hypothesis statements, producing an entailment score (“Score”) for each comparison. Some of the hypothesis statements are true based on information from the BSD (labeled “E” for “entailment”) and some are false (labeled “C” for “contradiction”). The maximum entailment score (MES) for true statements is compared to the MES for false statements to assess the quality of model inferences. Shown here is the process for a single registry field: site of surgery. The real experiments usually involved hundreds of premise sentences and dozens of hypothesis sentences based on 43 different registry fields. Note: The sentences shown here were invented for the purposes of this example and do not refer to any real patient.

In 2020, a team at Facebook released five pretrained NLI models with different transformer-based architectures **[9]**. Here we apply these models to curate a clinical registry for breast oncology, presenting a breakdown of model performance by registry field as well as a global comparison of models. Were this approach successful, it would allow us to build registries automatically from patient medical records, greatly increasing the speed and accuracy of curation and minimizing the effort required of human curators. It would also enable the rapid curation of registries for complex, rare, and underfunded diseases.

## Results

### Characteristics of patient population and documentation

A graphical summary of the study is shown in **Figure 2**. The study population consisted of 7439 patients who underwent surgery in the Mount Sinai Health System (New York, NY, USA) between January 1, 2010 and December 10, 2020 and had information recorded in an internal clinical research database (henceforth abbreviated as “BSD” for “Breast Surgery Database”), as well as one or more clinical, laboratory, or pathology notes recorded in the Mount Sinai Epic electronic health record (EHR) system within one year of surgery.

**Figure 2:**
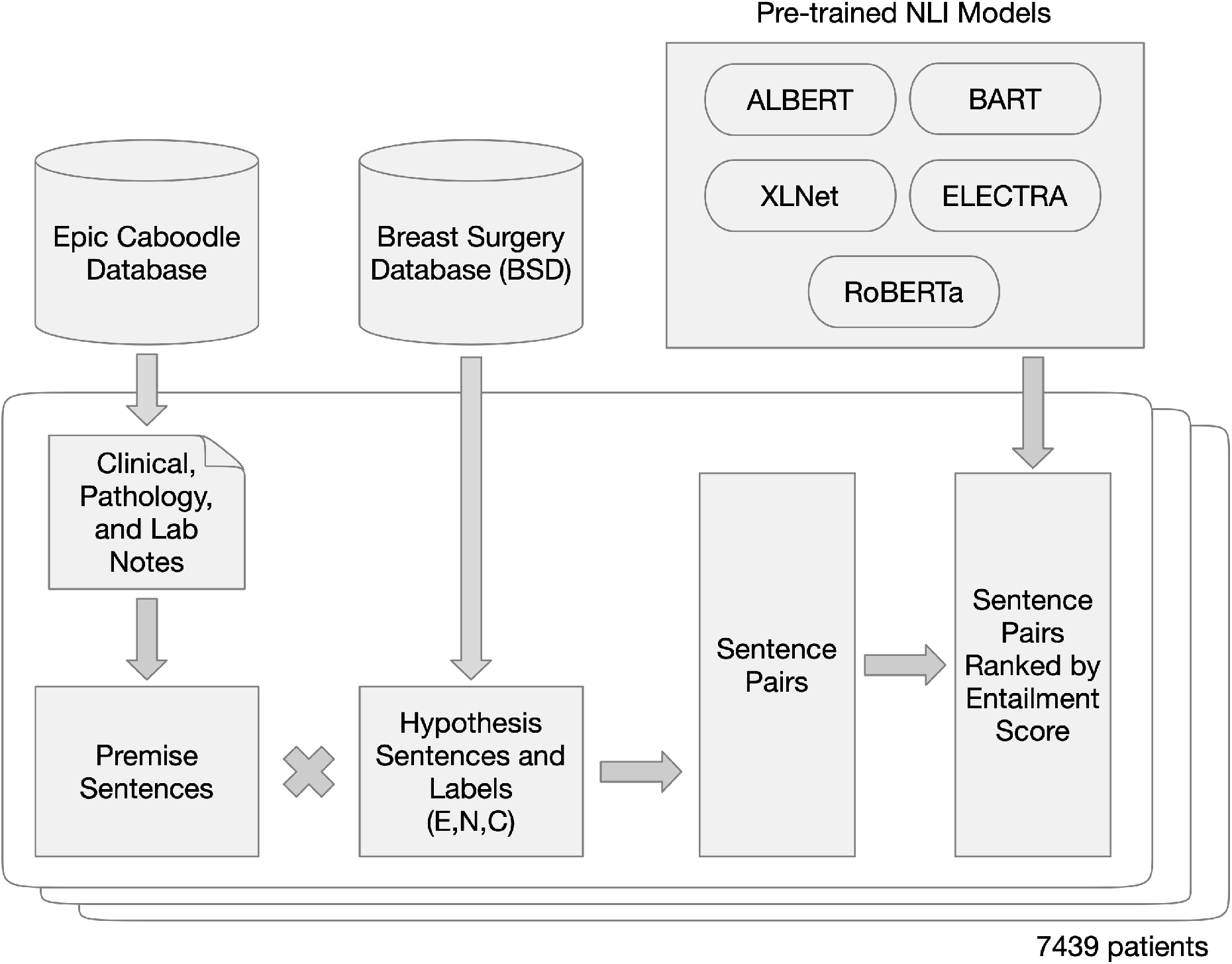
Graphical overview of the study. Premise sentences were extracted from clinical, pathology, and laboratory notes. Hypothesis sentences were generated from structured data in the BSD following the protocols in Tables 1 and 2. All possible combinations of premise and hypothesis were evaluated by one of five pre-trained NLI models. The top-ranked sentence for each hypothesis was extracted along with its entailment score. The process was repeated for all 7439 patients and all 5 NLI models.

Of the 7439 patients, 7386 (99.3%) were female and 53 (0.7%) were male. The median age at the time of surgery was 53.2 years (range: 13.0-97.5; IQR: 44.4-64.0). The patients’ self-reported races, in order of frequency, were White/Caucasian (4254; 57.2%), Other (1324; 17.8%), Black or African American (935; 12.6%), Asian (405; 5.4%), Pacific Islander (49; 0.7%), and American Indian or Alaska Native (4; <0.1%); for 468 patients (6.3%) the race was listed as Unknown or Not Reported. Ethnicity information was also self-reported; 4430 patients (59.5%) listed their ethnicity as Not Hispanic or Latino, 897 (12.1%) as Hispanic or Latino, and 2112 (28.4%) did not provide ethnicity information.

The number of unique sentences in documents recorded in clinical, pathology, and laboratory notes in Epic within one year of a patient’s surgery date ranged from 2 to 19835. The median number of unique sentences was 374 (IQR: 175-778). Thirty-two patients (0.4%) had more than 5000 unique sentences.

### NLI model accuracy and relative performance

True and false hypothesis statements were generated programmatically from structured information in the BSD using the strategies outlined in **Supplementary Tables 1 and 2**. The total number of premise-hypothesis comparisons made by the models was 521,449,945 (approximately 104 million per model x 5 models). Models were evaluated by their ability to score true inferences above false inferences across 43 different registry fields. A summary of the models’ performance is shown in **Table 1** and a breakdown by registry field is in **Table 2**.

**Table 1:** Summary of model performance. For each of the 43 possible hypothesis statement categories, the models were ordered according to the percent of patients for whom the maximum entailment score for true statements, MES-T, was greater than the MES for false statements, MES-F.

| Model | Overall (n=43) |  |  | No Laterality (n=17) |  |  | Laterality (n=26) |  |  |
| --- | --- | --- | --- | --- | --- | --- | --- | --- | --- |
|  | Rank Sum | Times Ranked 1st | Times Ranked 1st or 2nd | Rank Sum | Times Ranked 1st | Times Ranked 1st or 2nd | Rank Sum | Times Ranked 1st | Times Ranked 1st or 2nd |
| ALBERT | **90** | **22** | **28** | **42** | **8** | **10** | **48** | **14** | **18** |
| BART | 121 | 4 | 18 | 49 | 1 | 8 | 70 | 6 | 12 |
| RoBERTa | 126 | 9 | 17 | 50 | 2 | 5 | 71 | 2 | 13 |
| XLNet | 142 | 3 | 15 | 56 | 3 | 5 | 93 | 2 | 7 |
| ELECTRA | 166 | 5 | 8 | 58 | 3 | 6 | 108 | 2 | 2 |

**Table 2:** Model performance by hypothesis category. Hypothesis categories are ordered according to the median fraction of patients for whom the maximum entailment score for true statements, MES-T, was greater than the MES for false statements, MES-F.

|  |  |  |  | Fraction with MES-T > MES-F |  |  |  |  |
| --- | --- | --- | --- | --- | --- | --- | --- | --- |
|  | Hypothesis Category | Laterality | N | ALBERT | BART | ELECTRA | RoBERTa | XLNet |
| 1 | age |  | 7439 | 0.895 | 0.889 | 0.942 | **0.944** | 0.935 |
| 2 | site |  | 7439 | 0.933 | 0.922 | 0.912 | **0.938** | 0.934 |
| 3 | reconst |  | 7294 | **0.895** | 0.792 | 0.629 | 0.828 | 0.875 |
| 4 | neo_adj |  | 5185 | **0.929** | 0.735 | 0.370 | 0.643 | 0.831 |
| 5 | sln_bx_left | X | 2646 | 0.732 | 0.669 | 0.000 | 0.731 | **0.829** |
| 6 | palpable_right | X | 3955 | 0.670 | 0.700 | 0.695 | **0.753** | 0.562 |
| 7 | sln_bx_right | X | 2576 | 0.691 | 0.683 | 0.000 | 0.747 | **0.817** |
| 8 | path_dx |  | 7439 | 0.639 | 0.686 | **0.775** | 0.672 | 0.691 |
| 9 | palpable_left | X | 4096 | 0.656 | 0.711 | 0.663 | **0.735** | 0.605 |
| 10 | genetics |  | 7439 | 0.640 | 0.648 | **0.704** | 0.691 | 0.598 |
| 11 | fhx_breast |  | 7439 | 0.614 | **0.681** | 0.597 | 0.641 | 0.664 |
| 12 | gravida |  | 7439 | 0.638 | 0.599 | 0.561 | 0.446 | **0.671** |
| 13 | hormone_use |  | 7439 | **0.634** | 0.596 | 0.598 | 0.449 | 0.549 |
| 14 | smoking |  | 7439 | **0.799** | 0.771 | 0.569 | 0.498 | 0.587 |
| 15 | margins_right | X | 1786 | 0.516 | 0.516 | **0.536** | 0.410 | 0.209 |
| 16 | margins_left | X | 1893 | 0.499 | 0.549 | **0.556** | 0.443 | 0.225 |
| 17 | grade_left | X | 2123 | 0.496 | 0.559 | 0.426 | **0.563** | 0.432 |
| 18 | grade_right | X | 2024 | 0.495 | 0.506 | 0.428 | **0.533** | 0.344 |
| 19 | image_biop |  | 7439 | **0.690** | 0.467 | 0.675 | 0.169 | 0.116 |
| 20 | para |  | 7439 | 0.218 | **0.593** | 0.426 | 0.464 | 0.538 |
| 21 | lvi_right | X | 1627 | **0.536** | 0.307 | 0.451 | 0.458 | 0.487 |
| 22 | lvi_left | X | 1706 | **0.519** | 0.338 | 0.457 | 0.447 | 0.514 |
| 23 | status |  | 7439 | 0.423 | 0.316 | 0.346 | **0.518** | 0.378 |
| 24 | prin_finding_left | X | 4438 | 0.367 | 0.532 | 0.074 | **0.547** | 0.340 |
| 25 | prin_finding_right | X | 4281 | 0.364 | 0.517 | 0.075 | **0.555** | 0.349 |
| 26 | surgery_type_left | X | 7439 | **0.374** | 0.361 | 0.343 | 0.357 | 0.249 |
| 27 | ax_dissect_right | X | 2529 | **0.634** | 0.208 | 0.000 | 0.540 | 0.348 |
| 28 | fertility_rx |  | 7439 | **0.542** | 0.537 | 0.347 | 0.233 | 0.256 |
| 29 | sln_bx_left_n | X | 1769 | **0.499** | 0.383 | 0.291 | 0.343 | 0.239 |
| 30 | fhx_ovar |  | 7439 | **0.429** | 0.346 | 0.237 | 0.261 | 0.338 |
| 31 | surgery_type_right | X | 7439 | **0.350** | 0.336 | 0.327 | 0.346 | 0.227 |
| 32 | sln_bx_right_n | X | 1741 | **0.504** | 0.402 | 0.286 | 0.318 | 0.221 |
| 33 | ax_dissect_left | X | 2615 | **0.630** | 0.203 | 0.000 | 0.514 | 0.315 |
| 34 | prior_surg |  | 7439 | 0.209 | 0.226 | **0.266** | 0.212 | 0.228 |
| 35 | ax_dissect_right_n | X | 296 | 0.274 | **0.351** | 0.223 | 0.166 | 0.169 |
| 36 | personal_hx |  | 7439 | **0.208** | 0.205 | 0.131 | 0.207 | 0.192 |
| 37 | ax_dissect_left_n | X | 361 | 0.285 | **0.335** | 0.180 | 0.188 | 0.199 |
| 38 | er_right | X | 2211 | **0.269** | 0.191 | 0.029 | 0.114 | 0.200 |
| 39 | er_left | X | 2280 | **0.294** | 0.242 | 0.028 | 0.117 | 0.186 |
| 40 | her2_left | X | 1708 | **0.307** | 0.166 | 0.042 | 0.248 | 0.123 |
| 41 | her2_right | X | 1678 | **0.263** | 0.141 | 0.043 | 0.243 | 0.121 |
| 42 | pr_left | X | 2277 | **0.135** | 0.080 | 0.013 | 0.024 | 0.093 |
| 43 | pr_right | X | 2207 | **0.119** | 0.063 | 0.011 | 0.026 | 0.102 |

Five NLI models were evaluated: ALBERT **[10]**, BART **[11]**, RoBERTa **[13]**, XLNet **[14]**, and ELECTRA **[12]**. ALBERT outperformed the other models (scoring true inferences higher than false inferences for a greater fraction of patients) across 22 out of 43 (51.2%) registry fields. RoBERTa ranked second, coming in first in 9/43 fields, followed by ELECTRA (5/43), BART (4/43) and XLNet (3/43). Because the laterality (left or right breast) of certain categories was a potentially important confounder, we also considered model performance among only the 17 fields with no laterality. The ranking differed slightly: ALBERT again came in first, winning 8/17 fields, followed by XLNet and ELECTRA (tied for second with 3/17 fields each), RoBERTa (2/17), and BART (1/17).

There was considerable variation in model performance across registry fields. The registry field curated most easily by NLI models was patient age; RoBERTa, the best-performing model for that field, assigned a higher maximum entailment score (MES; see **Figure 1**) to true statements of age vs. false statements for 94.4% of patients. This was followed by site of surgery (RoBERTa; 93.8% correct), whether a breast reconstruction had been performed (ALBERT; 89.5% correct), and whether the patient had received neo-adjuvant therapy (ALBERT; 92.9% correct). The most difficult-to-curate fields were related to the estrogen receptor (ER), progesterone receptor (PR), and HER2 status of the breast masses. ALBERT outperformed the other models in all these cases. For ER status, ALBERT assigned a higher MES to true statements for 26.9% (right side) and 29.4% (left side) of patients; for PR, it was correct in 11.9% (right side) and 13.5% (left side) of patients; for HER-2, it was correct in 26.3% (right side) and 30.7% (left side) of patients.

There was also considerable variation in model performance across the five models within individual registry fields. For example, ELECTRA completely failed to identify true statements about whether sentinel node biopsies and axillary dissections were performed (**Table 2**, rows 5, 7, 27, and 33), even though the other four models performed well in these categories. ALBERT consistently outperformed the other models among the most difficult-to-infer fields (**Table 2**, rows 26 and below); for example, ALBERT was able to identify the correct number of sentinel lymph nodes biopsied in approximately 50% of cases, while performance among the other four models ranged from 22-40% (**Table 2**, rows 29 and 32). Among the “sided” fields, model rankings tended to be the same for both left and right breasts; for example, XLNet best identified whether a sentinel lymph node biopsy had been performed on the left side (82.9%; **Table 2**, row 5) and it was also the highest-ranking model for the same field on the right side (81.7%; **Table 2**, row 7).

### Error analysis and characteristic patterns

Examples of true and false hypothesis statements with high entailment scores, as well as the premise statements from patient EHRs that generated those scores, are shown in **Table 3**. A detailed error analysis by registry field is shown in **Table 4**.

**Table 3:**
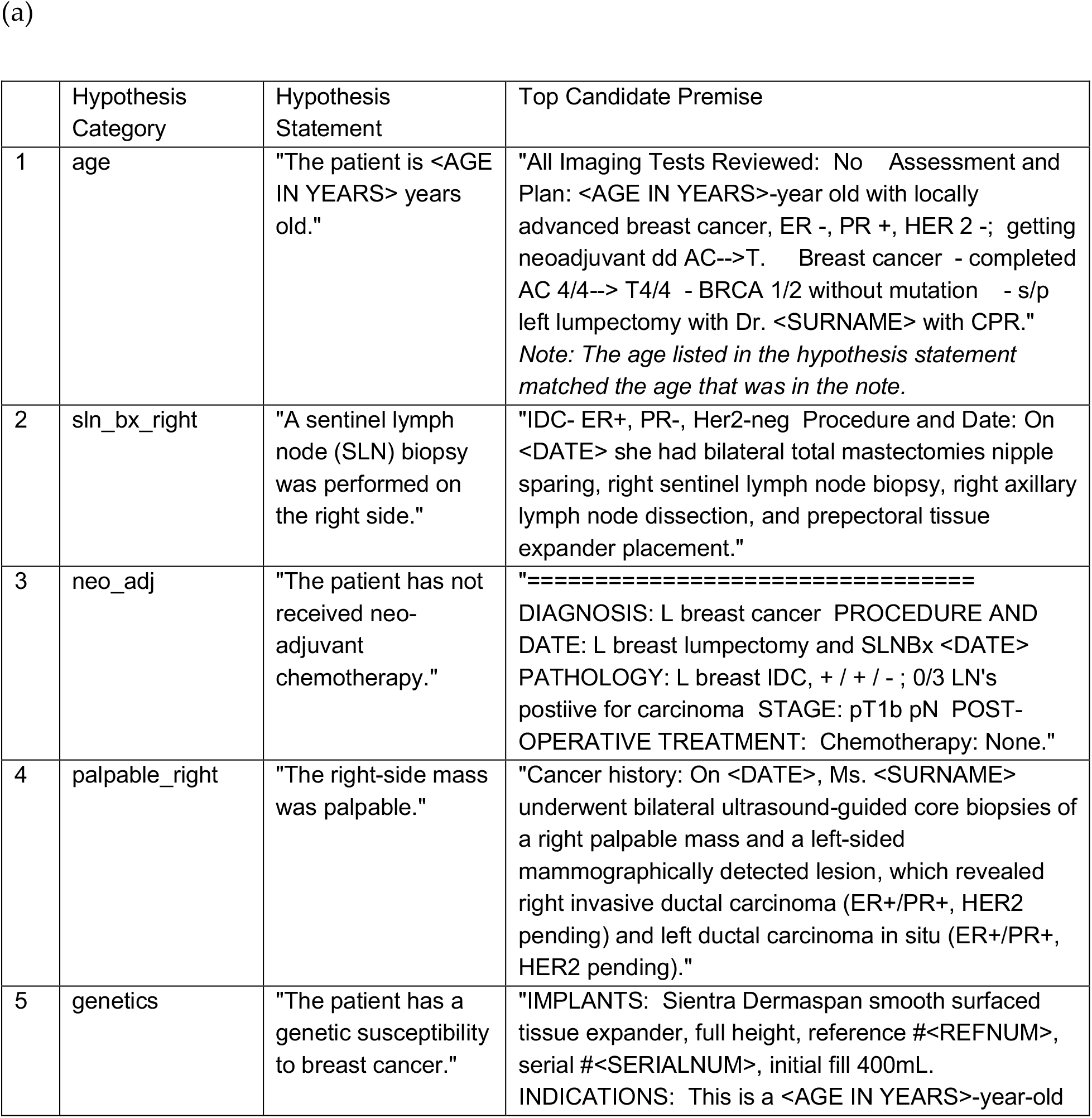
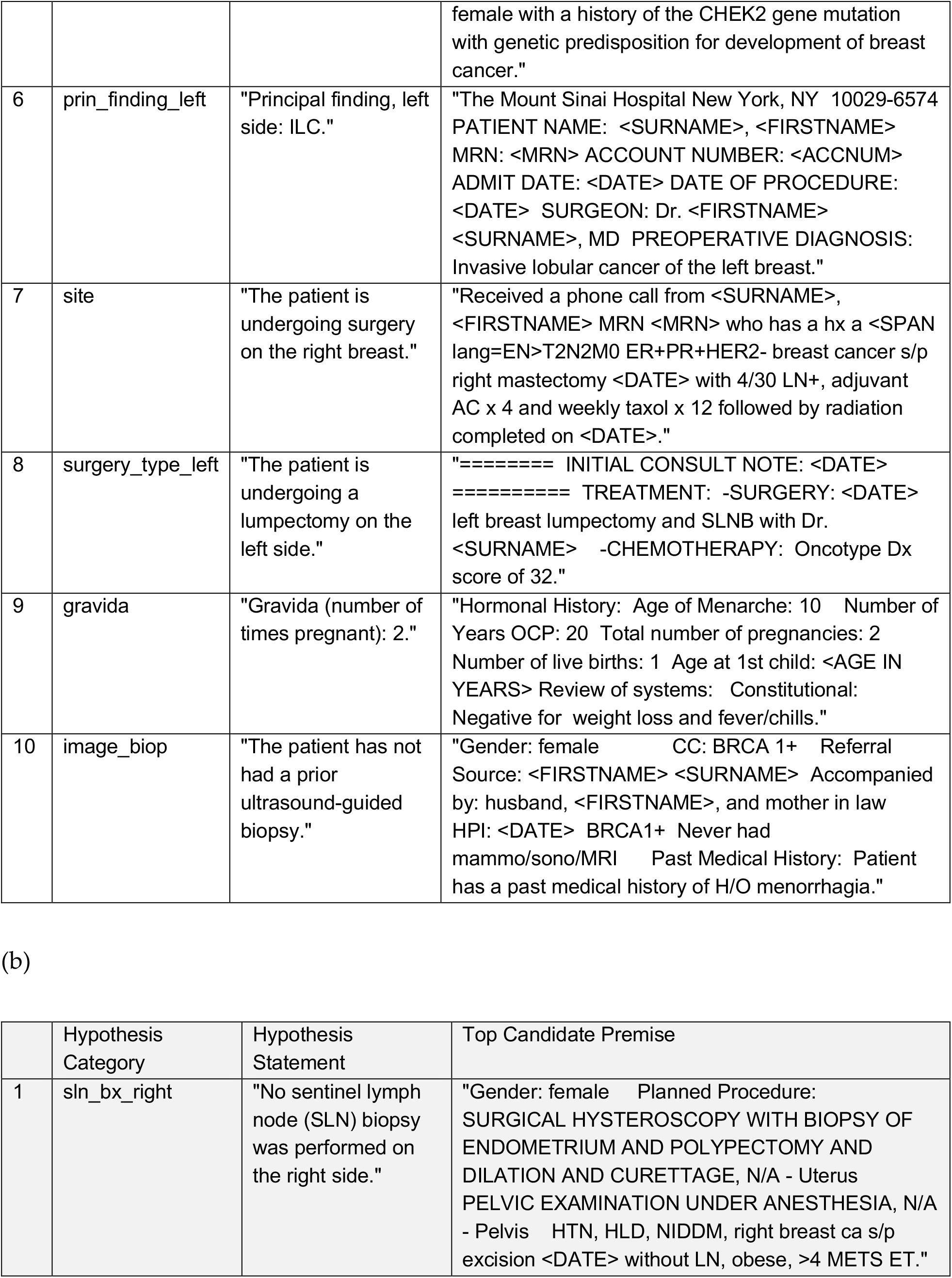
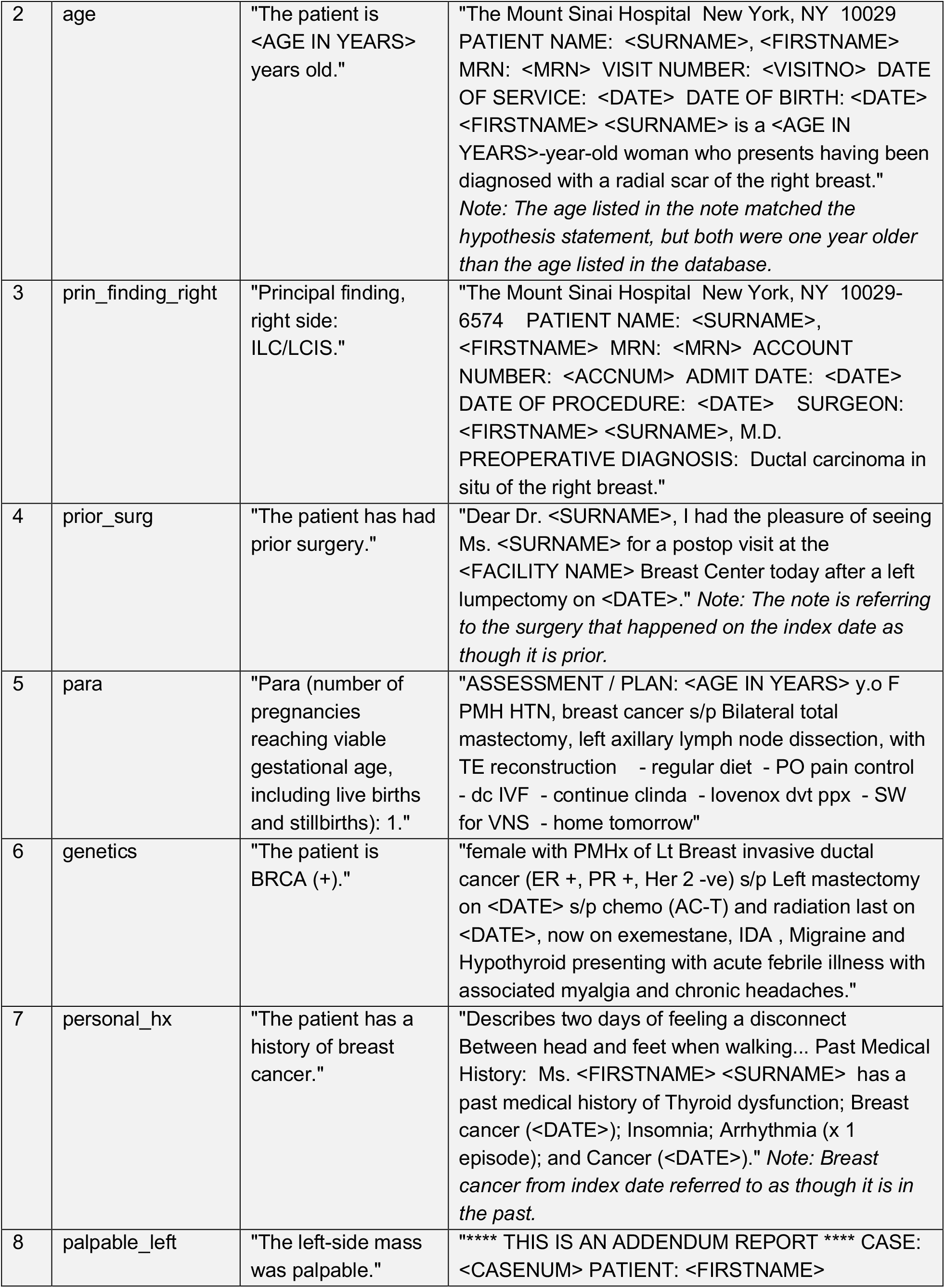
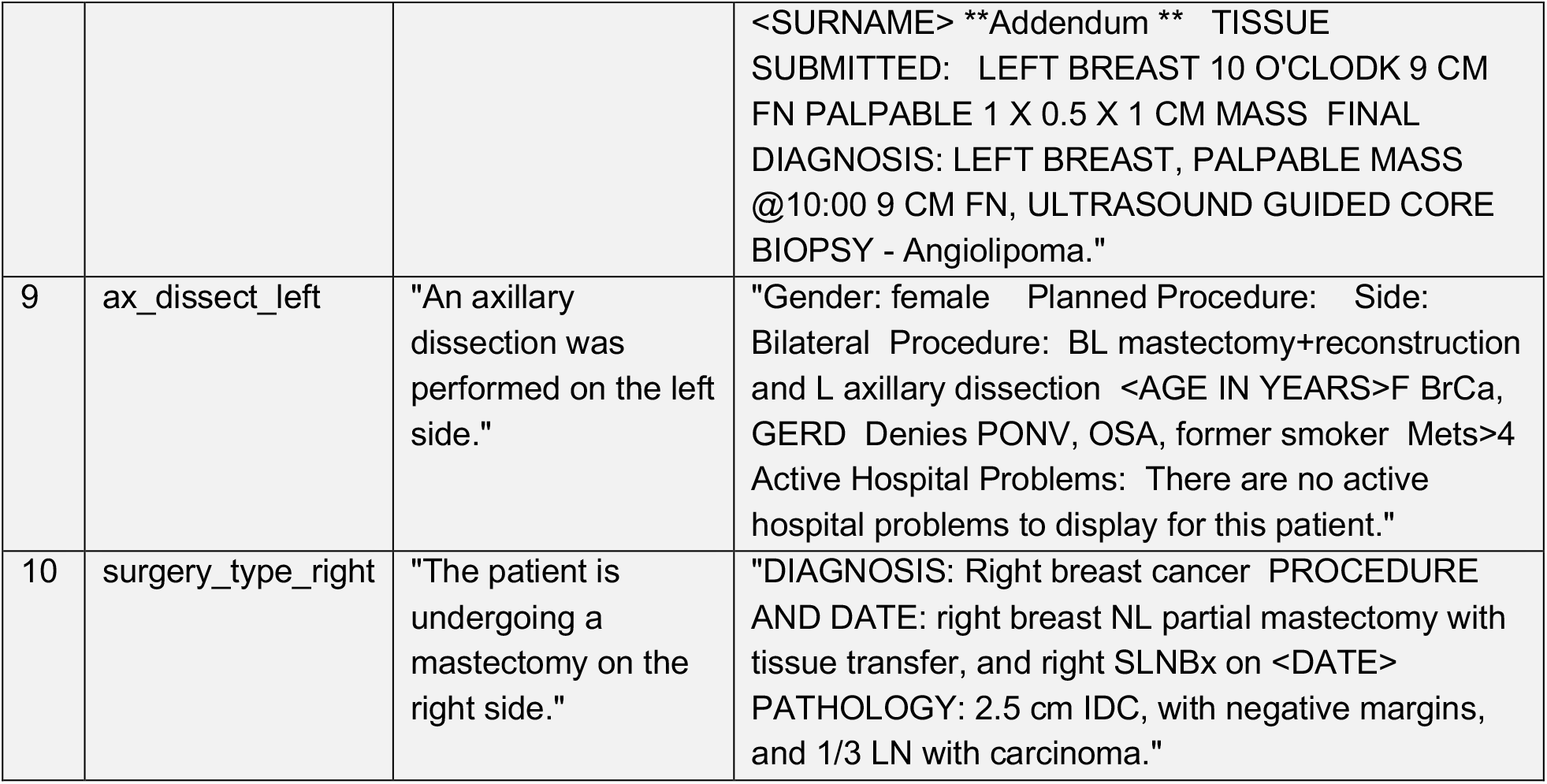
Top ten sentences with maximum entailment scores for the ALBERT model for hypothesis statements that were (a) true and (b) false based on information from the BSD. All entailment scores for the sentences shown here were greater than 0.99. To be included in the table, a sentence needed to be less than 300 characters in length. Only the top statement from each hypothesis category was included. All PHI elements and other potentially identifying information, such as physician names, were replaced by bracketed generic terms.

**Table 4:**
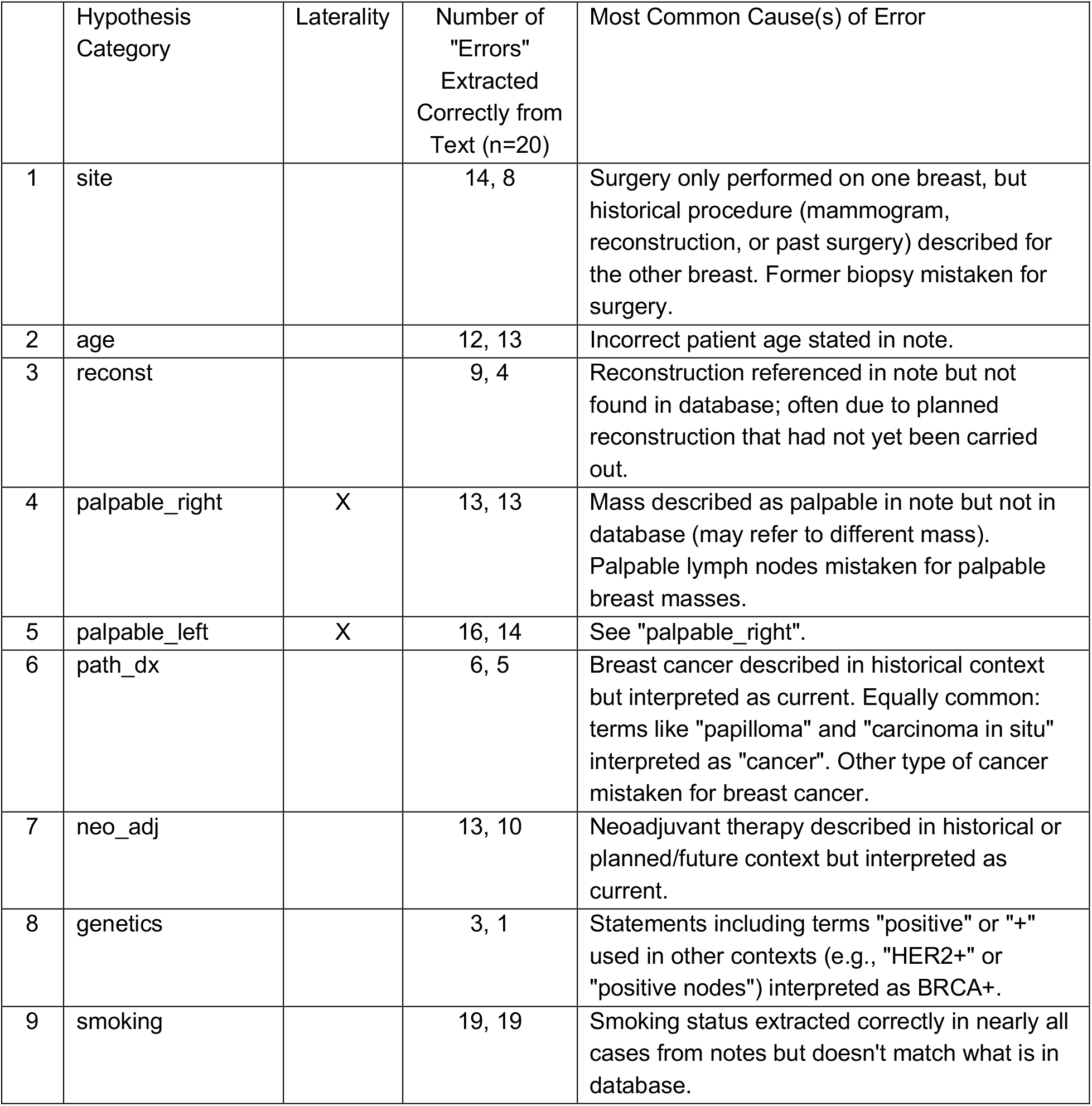
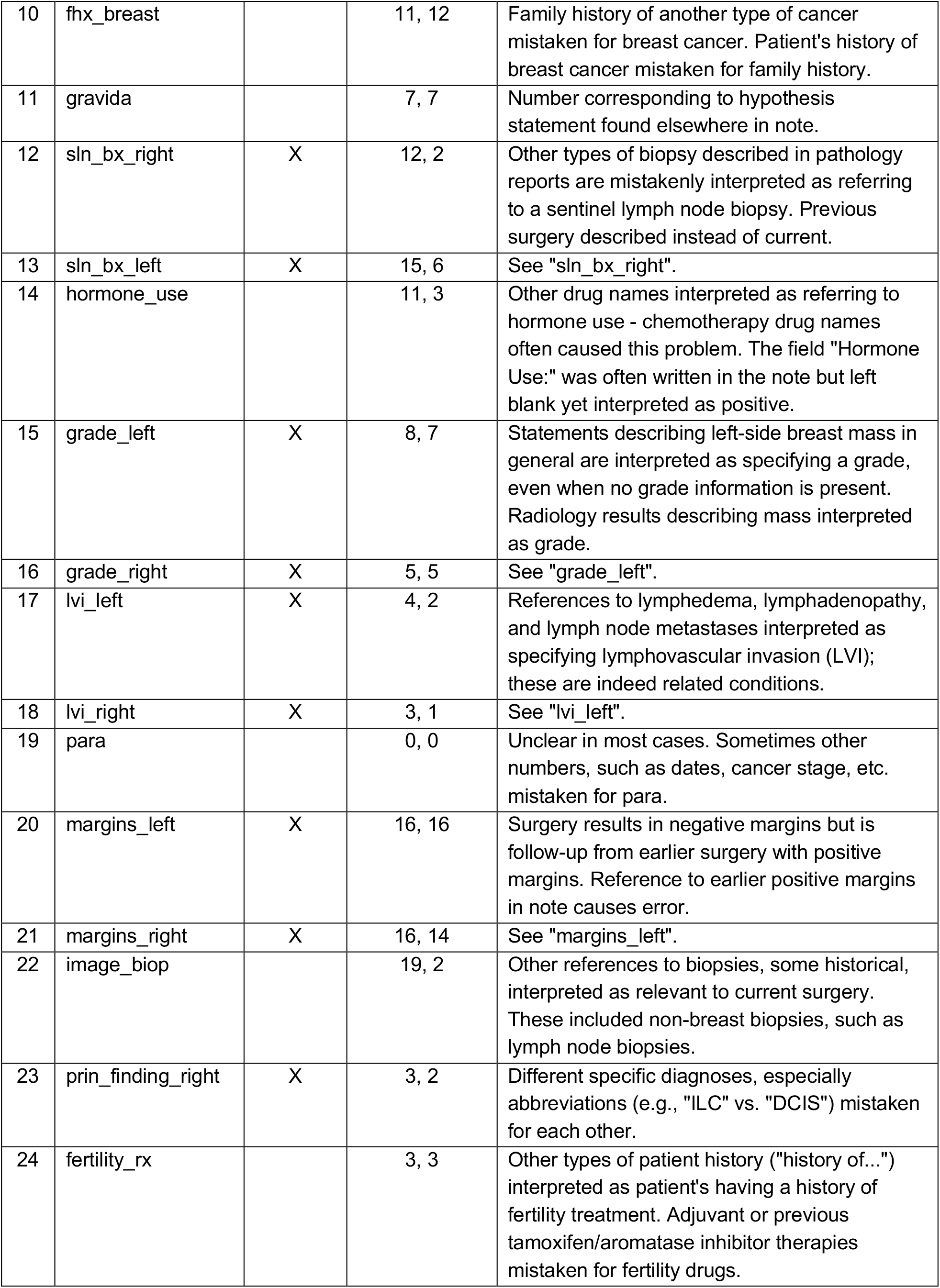
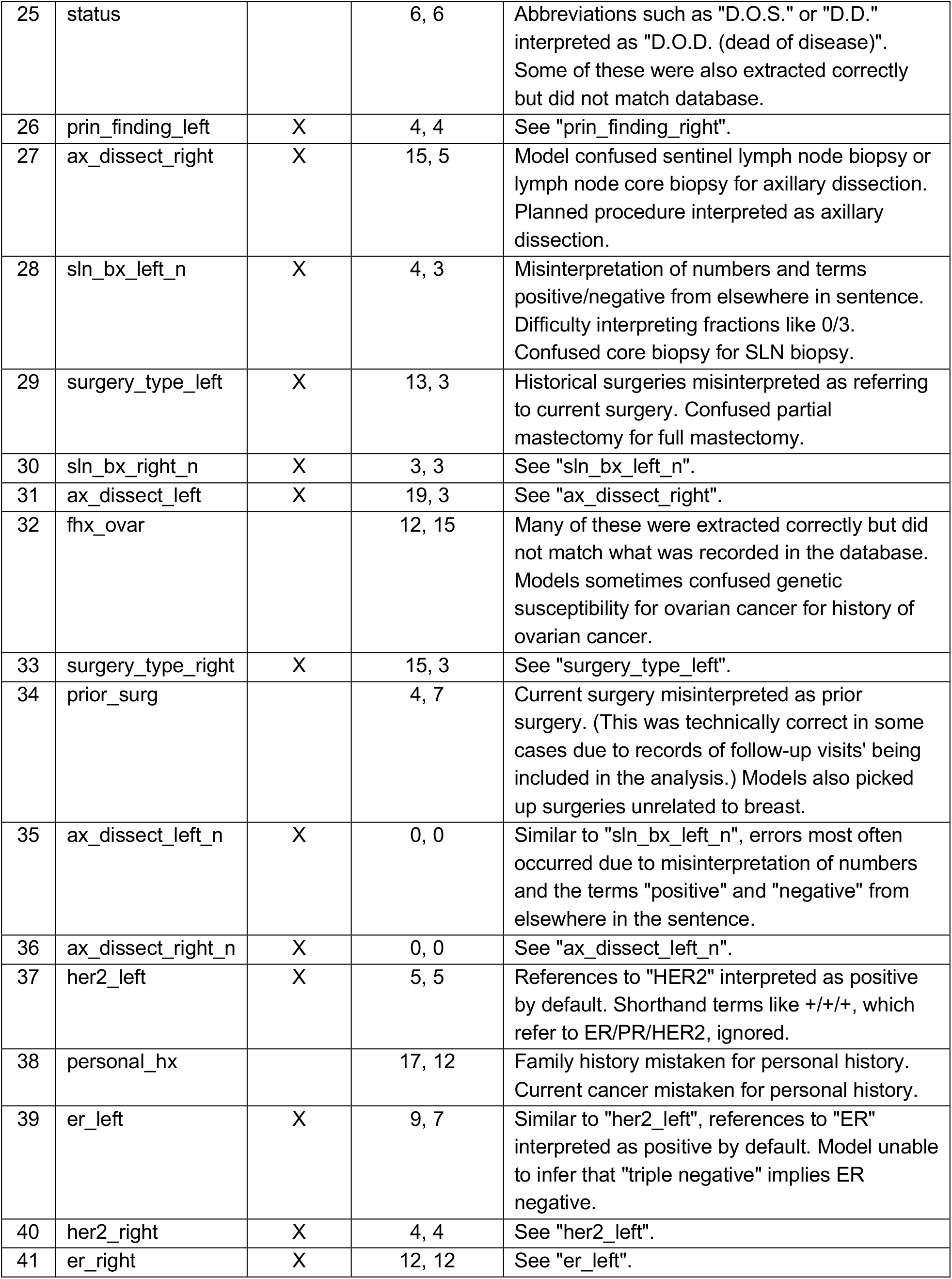
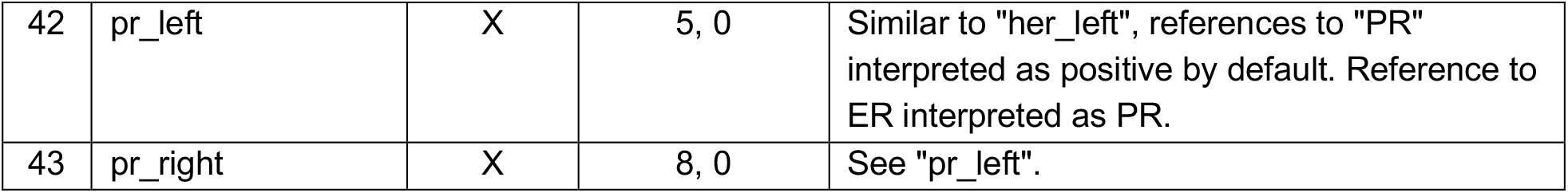
Top causes of error for each hypothesis category. The top 20 error sentences from each category (defined as sentences for which the MES was high but the hypothesis was false) were examined by two different reviewers to establish the most common cause(s) of error. Reviewer 1 (CG) was knowledgeable about the project but had no specific training in breast oncology. Reviewer 2 (KP) had a deeper knowledge of breast oncology and was part of the team that curated the BSD.

The most common errors arose from models’ misinterpretations of temporality; they interpreted prior or planned surgeries as current and vice versa. In part, this was a product of our experimental design; since notes were extracted from within a year of surgery, descriptions of the index surgery from before or after it occurred were almost always included. For example, a patient may have had no prior surgery on the index date (and hence be listed in the BSD as having had no prior surgery) but the surgery performed on the index date could be referred to as a “prior surgery” in a note written shortly thereafter. Manual review (**Table 4**) revealed that approximately a quarter of all “errors” --statements with high entailment scores that did not match the BSD--were indeed correct inferences based on what was stated explicitly in the text.

A second set of errors arose from models’ tendency to fixate on certain clinical terms. For example, the models often failed to distinguish between references to “palpable” breast masses and “palpable” lymph nodes (**Table 4**, rows 4-5); they also tended to interpret any reference to “biopsy” as an indication that a patient had undergone a sentinel lymph node biopsy (**Table 4**, rows 12-13). Lymphovascular invasion was often inferred incorrectly because notes contained references to other words beginning with “lymph”, such as lymphedema, lymphadenopathy, and lymphoma (**Table 4**, rows 17-18). Clinical terms like “papilloma” or “fibroadenoma”, which share a suffix with “carcinoma” (cancer), were also sometimes misinterpreted as indicating a diagnosis of breast cancer (**Table 4**, row 6).

Finally, there were a few cases in which what was written in the note directly contradicted the database, most likely because notes contained text that was copy/pasted over from earlier notes. One of these was age: patients’ age as written in notes sometimes did not match what one would obtain by subtracting the patient’s birth date from the date the note was written (**Table 4**, row 2). The patient might be described as a “30-year-old female” but be 32 years of age, for instance. A second case related to smoking status. Over half the patients in the BSD were listed as nonsmokers at the time of surgery (or had no smoking status listed) but the NLI models sometimes picked up instances where a supposedly nonsmoking patient was described as a former or occasional smoker in notes (**Table 4**, row 9).

## Discussion

Our goals in this project were (1) to establish a baseline for NLI performance in registry curation and (2) to uncover characteristic error patterns that could help guide the development of future NLI models. Hundreds of papers have applied NLP to clinical text **[6]**, but most of these approaches are inappropriate for the registry curation problem, either because they rely on the existence of specially annotated training data or because they would necessitate building a separate model for each registry field. NLI presents a possible path forward.

### Factors affecting NLI model performance

Although the five NLI models shared many architectural similarities, the ALBERT-based model outperformed the others on a majority (22 out of 43) registry fields. The details of the ALBERT model offer clues to its enhanced performance: it incorporates factorized embeddings and cross-layer parameter sharing, which reduce the total number of parameters learned by the model **[10]**. These choices were designed to permit pretraining on much larger datasets: in other words, ALBERT sees much more data during pretraining for a similar amount of computational cost. This appears to provide the model with a significant performance lift on the registry curation task and suggests that the size of the pretraining corpus is a significant factor in clinical NLI.

A second important consideration is domain specificity. We were initially concerned that the high frequency of specialized terms and abbreviations in clinical text would cause the pretrained NLI models to fail e.g., select random and irrelevant premise statements and essentially act as random number generators. However, this turned out not to be the case. The models, which were trained using general-domain corpora (text from news articles, blogs, books, Wikipedia, etc.), made correct inferences for at least 50% of patients on over half of the registry fields tested (**Table 2**). This observation has important implications for the focus of future clinical NLI model development. For example, to date there have been a few attempts to pretrain BERT on clinical text **[24, 25]** and create NLI training sets specific to clinical text **[23]**. However, the corpora involved in these studies are orders of magnitude smaller than the general-domain corpora used to pretrain and fine-tune the models used in our experiments **[9]**. If corpus size is the determining factor in the quality of NLI models for registry curation, it may make sense to combine corpora or NLI training sets from the clinical and general domains, rather than train clinically specific models.

Finally, we observed that even when the NLI models made incorrect inferences, they almost always chose premise sentences from the patient’s record that contained the information needed to determine the truth or falsehood of a given hypothesis (**Table 3**). This suggests that even if NLI models are not sufficient for end-to-end registry curation, they could be effective in software that filters patient records to identify relevant sentences and paragraphs. Whether this could also be accomplished through simpler methods like term matching remains to be seen.

### Computational considerations and model deployment

Modern NLP methods based on pretrained transformer models are extremely computationally expensive. The pretraining process for BERT **[21]** and related architectures is so resource-intensive that most active development is happening at large technology companies, such as Facebook and Google, and well-funded research labs at institutions like OpenAI and Stanford. Other groups wishing to use these models typically apply a transfer learning approach **[22]**, in which pretrained models are downloaded and then fine-tuned to perform downstream tasks, such as NLI. Even the fine-tuning step is likely to be prohibitive for most healthcare institutions, however; most will lack access to GPUs and the expertise needed to train and deploy these models. For this reason, we opted to deploy the five pretrained NLI models “out of the box”, even though they had been fine-tuned on general-purpose NLI corpora (SNLI, MNLI, FEVER, and ANLI) and might have benefited from further training on a clinical NLI corpus like MedNLI **[23]**. Although we performed no additional training, even the inference process was expensive on our 7439-patient dataset: it took approximately one month to apply the five pretrained models to all ∼521 million premise-hypothesis pairs in our experiments, running between 10 and 30 jobs in parallel at any given time. Of course, most registries are built over a period of years, and there would rarely be a need to batch process an entire registry at once as we did.

Deploying an NLI model for registry curation requires the thoughtful design of hypothesis statements for all of the different registry fields, as we have outlined in **Supplemental Tables 1 and 2**. This process takes time and should be checked by a domain expert. Ideally, multiple forms of each hypothesis would be included to reduce the impact of noise and/or models’ fixation on particular terms.

### Study limitations

Our study faced several data extraction and preprocessing challenges, our responses to which may have affected model performance and/or limited the applicability of these methods. First, the NLI models we used can only handle single-sentence inference, meaning that both premise and hypothesis can only be single sentences. More complex patterns of reasoning, e.g., by combining information from multiple notes in a patient’s record, would require different methods. Second, because the patients in the BSD had surgery at different times over a span of a decade, they had different amounts of text available in Epic. Through our experimental design, we were effectively assuming that the information needed to make inferences about all our BSD-derived hypothesis statements was present in the text extracts we obtained from Epic; in reality, this was not guaranteed to be the case. Because our results somewhat conflate data availability and model performance, we present a detailed breakdown of model performance by patient data availability in **Supplementary Table 3**. Finally, the models’ struggles with temporal reasoning might have been alleviated by applying heuristics or treating the different registry fields differently. Information about different registry fields can be found in multiple locations within the patient’s EHR, and some registry fields are more likely to be found in specific places - pathology reports, post-operative reports, etc. We could have applied restrictions to the models to enable them to access text from only these sources. We could likewise have applied date restrictions, allowing certain fields access to a narrower set of notes than others to avoid problems stemming from confusion of past, present, and planned surgeries. Since this study was designed to establish a baseline for NLI performance on this task, however, we chose to employ the simplest approach possible (allowing all fields access to all notes) even if it hurt model performance.

### Conclusions and future work

As the volume of EHR data in the United States and around the world grows **[26]**, we are in increasingly greater need of systems that can structure and curate this information automatically. Clinical registries are a fundamental component of high-quality clinical research and operational planning, yet the quality and timeliness of registry data still hinge on human curators’ ability to interpret vast amounts of clinical text. This is exactly the sort of detailed, repetitive task that most benefits from computational approaches. However, the structure of the task, particularly the need to curate dozens of different registry fields simultaneously, puts it beyond the reach of today’s most popular clinical NLP systems, most of which are limited to simpler tasks like named entity recognition and text classification **[6, 19, 27]**.

This study is, to our knowledge, the first to deploy modern NLI methods on a practical, clinical task that is not part of a computer science shared task or benchmark. We hope it provides encouragement to the vibrant and growing NLI community to continue to develop NLI technology in a way that will permit healthcare institutions--even those without computer science expertise or sophisticated computational resources--to use NLI for registry curation and related problems that directly impact patient lives.

## Methods

### Data access, storage, and processing

Raw clinical, laboratory, and pathology notes were obtained by querying the Mount Sinai Epic Caboodle database (Epic Systems Corp., Verona, WI, USA). Notes were included if they were written within one year of a patient’s surgery date, as recorded in the BSD. Notes were broken into sentences using the spaCy **[19]** default NLP pipeline. The BSD itself is stored in a RedCap database **[1]** and was obtained via direct export. If a patient had more than one surgery, we restricted our study to information from the first surgery date. This was there is no reason why the methods described here could not be applied to all surgeries, but we did not want patients with multiple surgeries to dominate the evaluation.

This study was approved by the Mount Sinai Institutional Review Board. Hypothesis statement generation and interpretation of results were overseen by a breast surgery attending (HS). All data storage and analysis were performed on Minerva, the Icahn School of Medicine’s HIPAA-compliant supercomputing cluster. This project benefited from our use of Minerva’s NVIDIA V100 GPU compute nodes, which greatly sped up the computations required to apply the pretrained NLI models to hundreds of millions of sentence pairs.

### Translation of structured database fields into hypothesis statements

A full description of how hypothesis statements were generated and labeled for each patient based on BSD information can be found in **Supplementary Tables 1 and 2**. For numeric fields, like age, multiple statements were generated, only one of which contained the correct age; typically, 4-5 false statements were generated for each true statement. For categorical fields with few levels, like smoking status, one statement was generated for each level (e.g., “current”, “former”, “never”, “unknown”), only one of which contained the correct value. For string fields, such as principal finding, which could have hundreds of levels, 4-5 strings randomly sampled from the rest of the database were used to create false statements, while the patient’s true string was used to generate a true statement.

### Application of pre-trained NLI models

Five pre-trained NLI models were used. We denote each by the name of its pretraining approach: ALBERT **[10]**, BART **[11]**, ELECTRA **[12]**, RoBERTa **[13]**, and XLNet **[14]**. These models were released by a team at Facebook in 2020 **[9]** and stored on the HuggingFace **[15]** model repository. All had been fine-tuned for NLI using a combination of NLI datasets, including SNLI **[16]**, MNLI **[17]**, FEVER-NLI **[18]**, and ANLI (R1, R2, R3) **[9]**.

After converting the structured information from the BSD into hypothesis statements for each patient, we applied these pretrained NLI models to the Cartesian product of hypothesis statements and all “premise” sentences from the patient’s clinical, laboratory, and pathology notes. Each of these comparisons produced an entailment score: the model’s level of certainty that the premise implied the hypothesis. The maximum entailment score (MES) was, for each hypothesis statement, its highest entailment score across all possible premise statements for that patient. This approach also identified the specific premise statement that produced the MES.

Our measure of performance for the NLI models was the fraction of patients for whom the MES for true statements (MES-T) was higher than the MES for false statements (MES-F) for statements pertaining to a particular registry field. We summarized this measure for each model-field combination (**Table 4**) and created a global summary of each model’s performance across fields (**Table 3**).

## Data Availability

The breast surgery database and raw clinical notes used in this study contain PHI and cannot be made available except through special arrangement via the Mount Sinai Data Use Committee. Please contact the corresponding author with any inquiries.

## Acknowledgments

This work was supported in part through the computational resources and staff expertise provided by Scientific Computing at the Icahn School of Medicine at Mount Sinai. We would particularly like to thank Lili Gai, who provided extensive assistance with the Minerva GPU nodes and helped troubleshoot several failed job submissions.

## Author Contributions

KP and HS created the initial problem specification for the project. BP chose NLI as a potential solution, designed and built the software for registry and note preprocessing and NLI model application, ran the NLI jobs on Minerva, and drafted the paper. CG extracted the raw clinical, laboratory, and pathology notes from Epic. CG and KP manually evaluated 860 sentence pairs each for the error analysis in Table 4. HS oversaw the project, provided technical advice and feedback, quality checked the auto-generated hypothesis statements, and served as our source of access to the BSD. All of the authors reviewed, edited, and proofread the final manuscript.

## Supplementary Information

**Supplementary Table 1:**
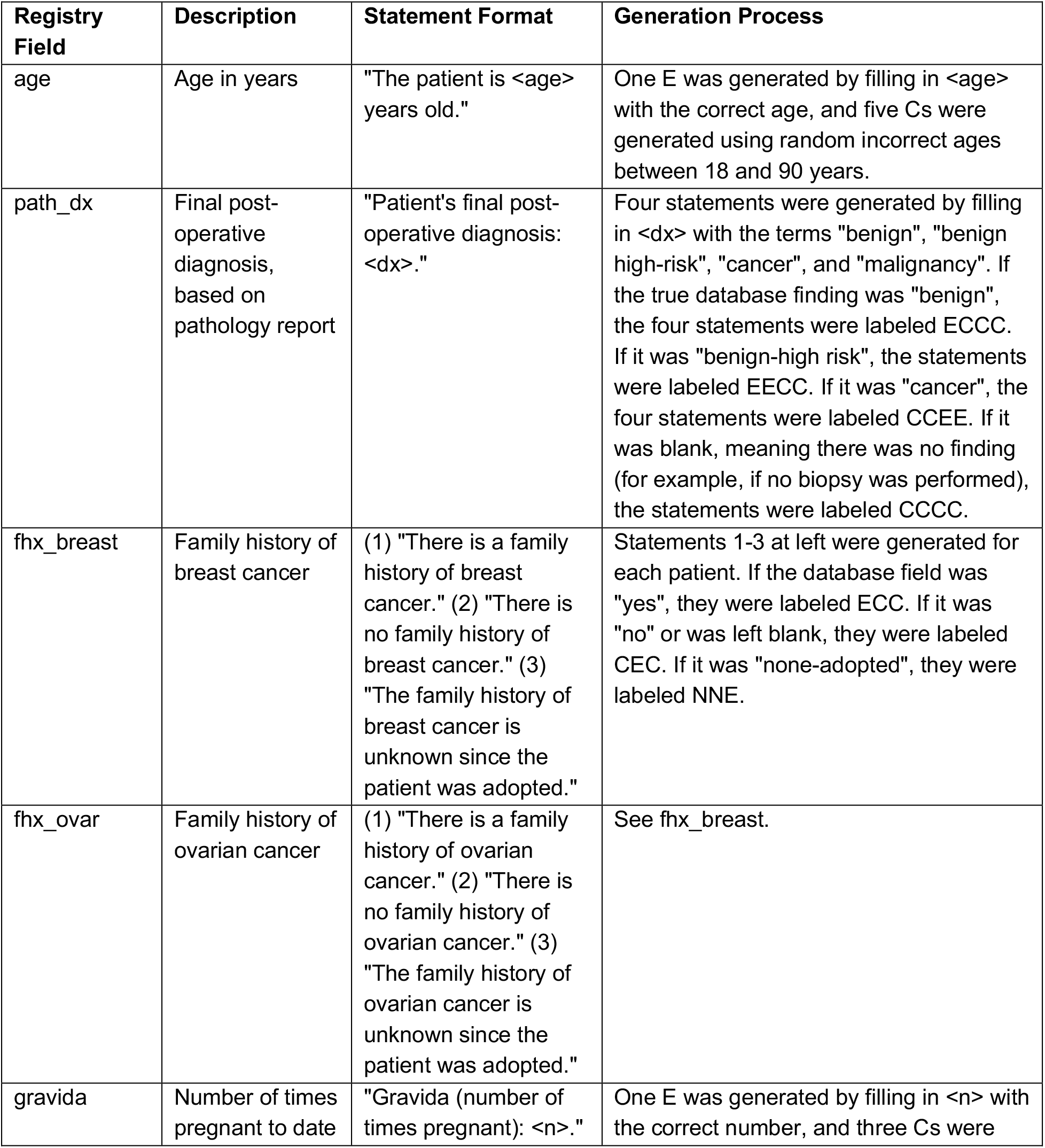
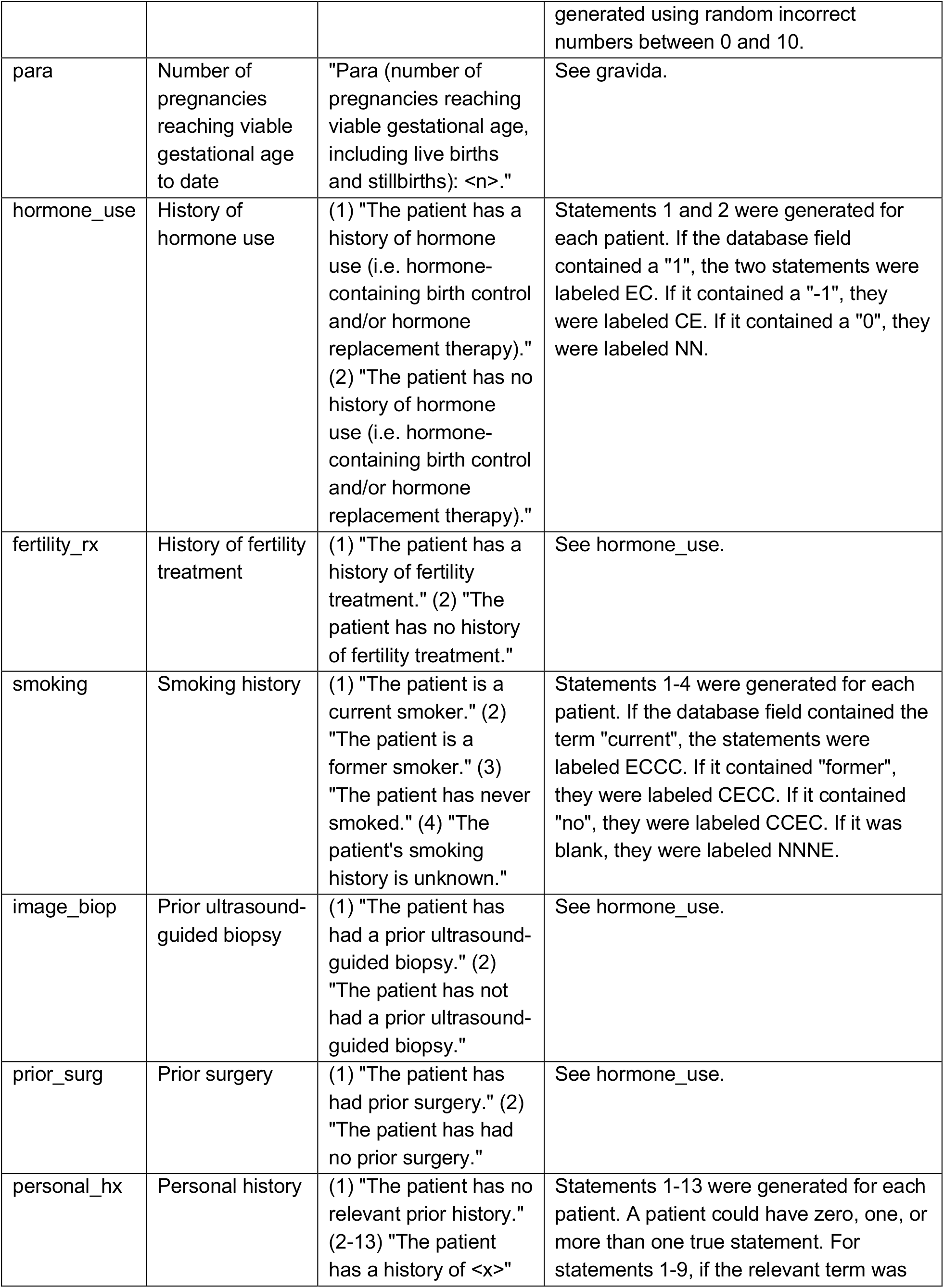
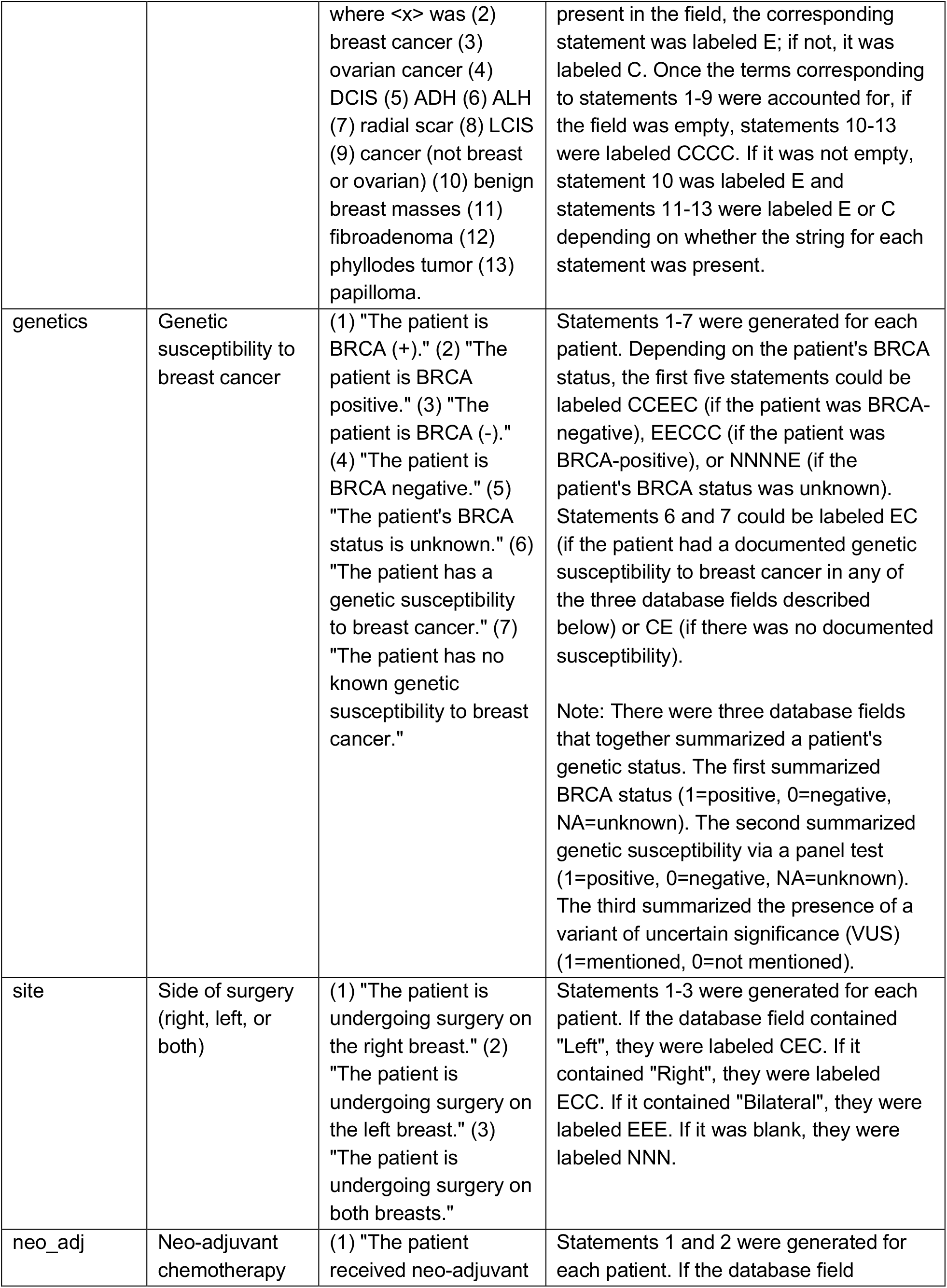
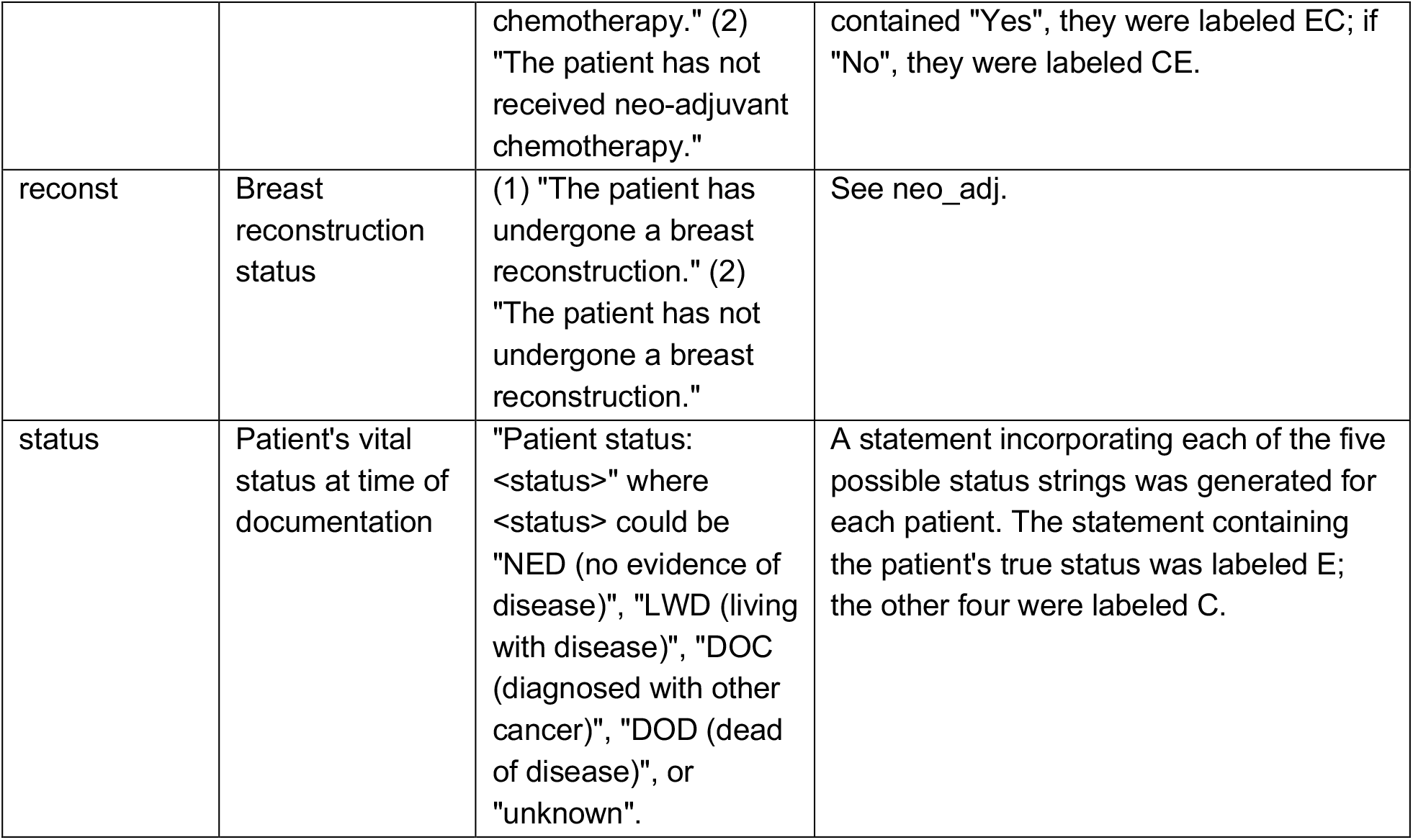
Generation of hypothesis statements based on structured registry data, Part 1 (statements referring to the whole patient). The abbreviation “E” refers to an entailed statement, “C” refers to a contradictory statement, and “N” refers to a neutral statement.

**Supplementary Table 2:**
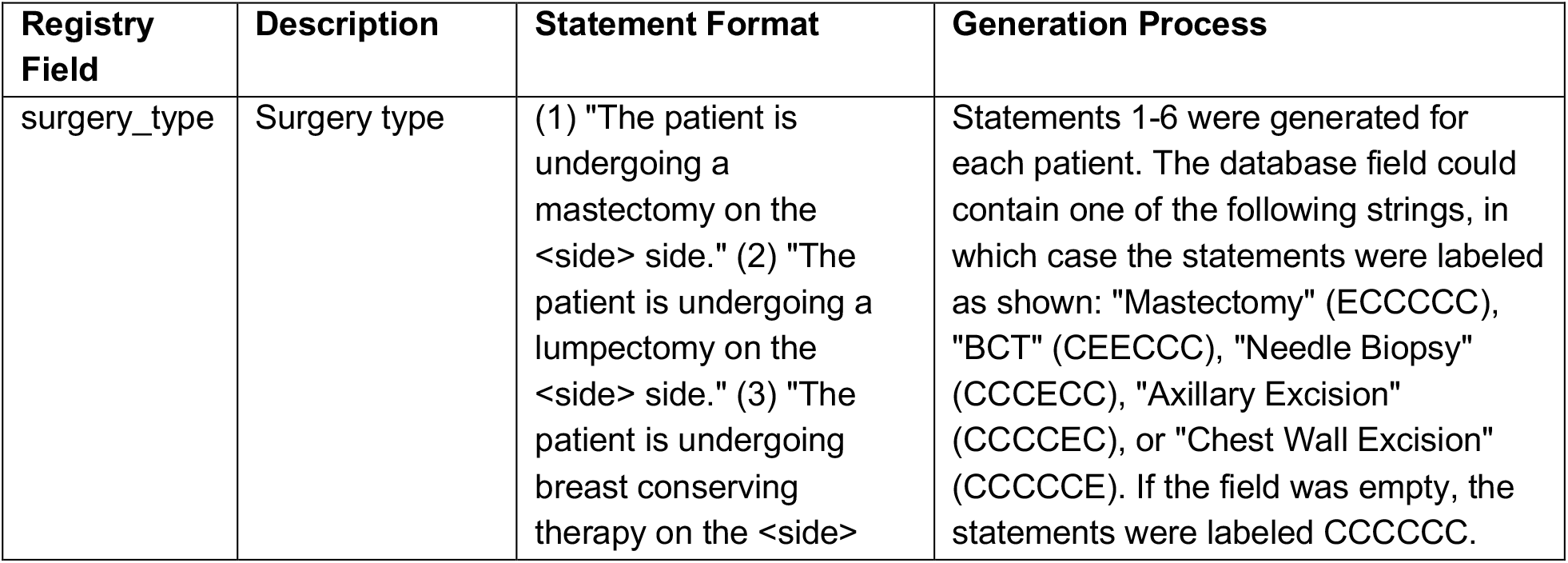
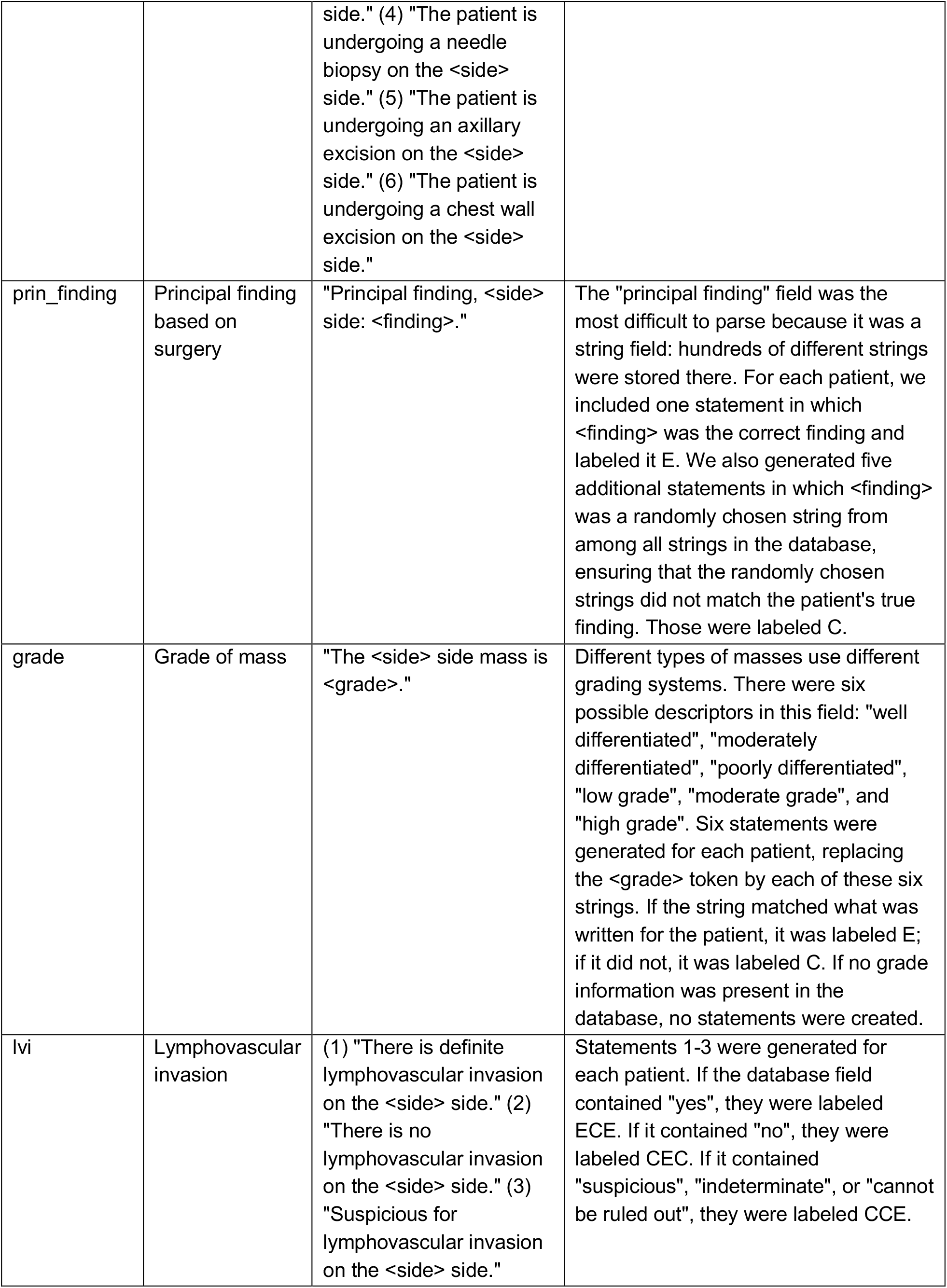
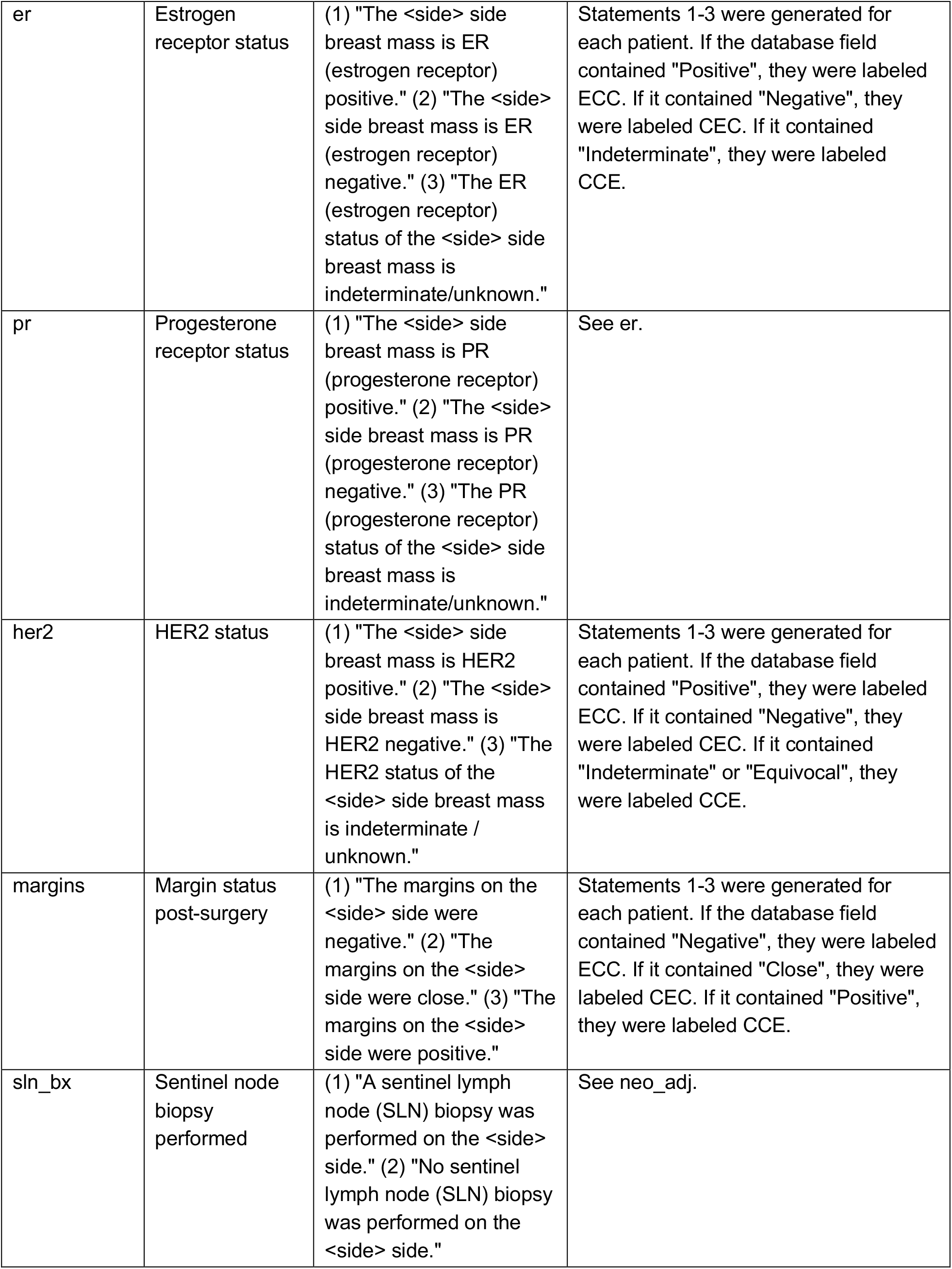
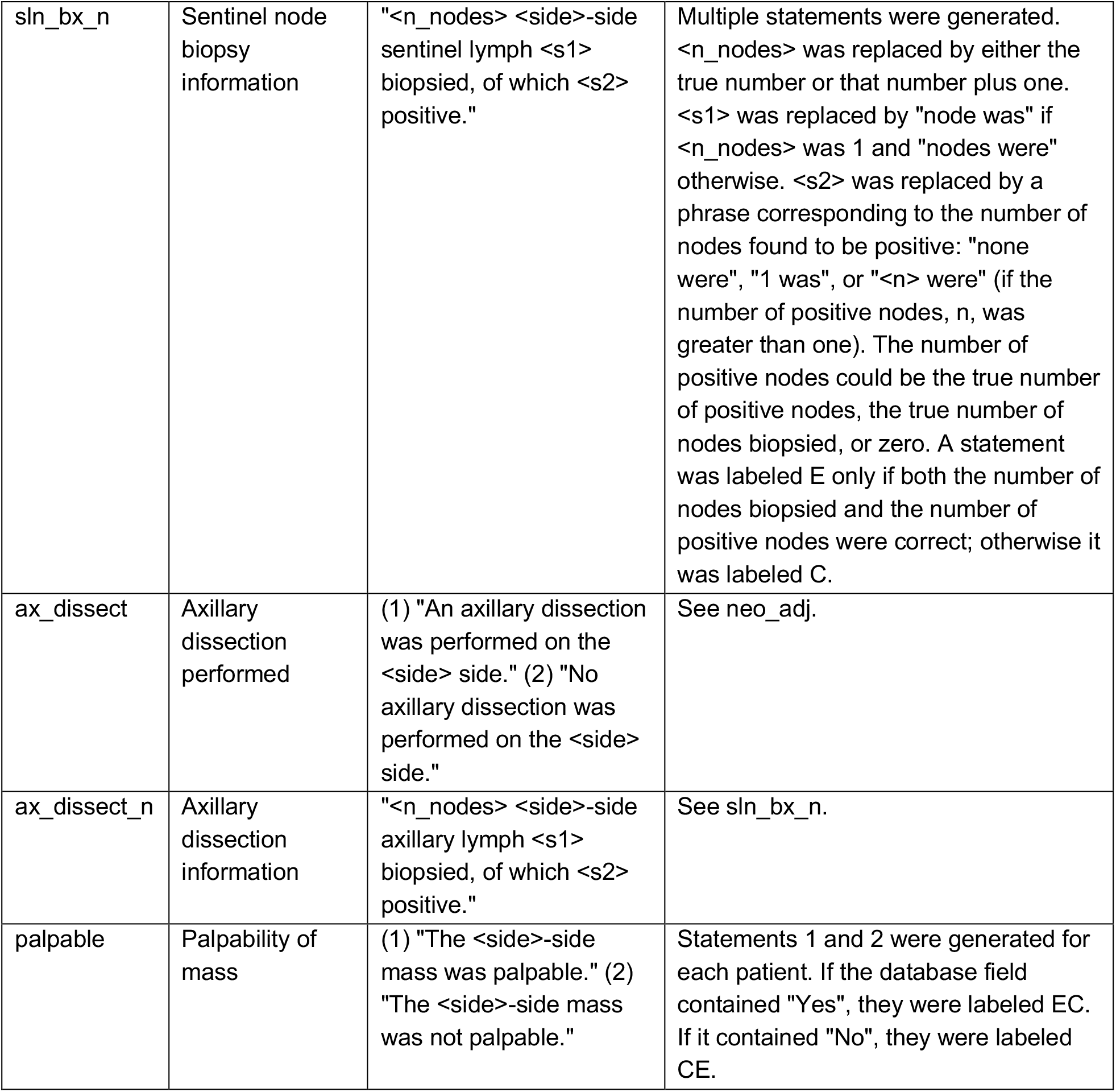
Generation of hypothesis statements based on structured registry data, Part 2 (statements generated separately for the left and right breasts). The variable <side> was filled in with either “left” or “right” . Information from the database field for “site” (Table 1) was used to determine which statements to generate. If a patient was having surgery on both sides, statements about both sides were generated. Abbreviations: “E” refers to an entailed statement, “C” refers to a contradictory statement, and “N” refers to a neutral statement.

**Supplementary Table 3:** Dependence of model performance on availability of patient data. Patients were divided into three groups: those with fewer than 100 sentences, those with 100-1000 sentences, and those with more than 1000 sentences.

(Full table available in attached csv file.)

